# Attention improvement to transcranial alternating current stimulation at gamma frequency over the right frontoparietal network: a preliminary report

**DOI:** 10.1101/2024.06.10.24308704

**Authors:** Tien-Wen Lee, Sergio Almeida, Gerald Tramontano

**Affiliations:** The NeuroCognitive Institute (NCI) Clinical Research Foundation, NJ 07856, US

**Author notes:** Corresponding author at The NCI Clinical Research Foundation, New Jersey, US Address: 111 Howard Blvd., Suite 204, Mt. Arlington, NJ 07856, Web: http://neuroci.com. Note: TW Lee and S Almeida contributed equally to this research as co-first authors. Email addresses for the other author: TW Lee; S Almeida.

**Keywords:** Attention, Transcranial alternating current stimulation (tACS), Transcranial electrical stimulation (tES), Frontoparietal network, Test of Variables of Attention (TOVA)

## Abstract

**Objective:** Applying transcranial alternating current stimulation (tACS) at 40 Hz to the frontal and parietal regions can improve cognitive dysfunctions. This study aimed to explore the influence of tACS at gamma frequency over right fronto-parietal (FP) region on attention.

**Methods:** We administered Test of Variables of Attention (TOVA; visual mode) to 44 participants with various neuropsychiatric diagnoses before and after 12 sessions of tACS treatment. Alternating currents at 2.0 mA were delivered to the electrode positions F4 and P4, following the 10-20 EEG convention, for 20 minutes in each session.

**Results:** We observed significant improvement across 3 indices of the TOVA, including reduction of variability in reaction time (RT; *P*=0.0002), increase in d-Prime (separability of targets and non-targets; *P*=0.0157), and decrease in commission error rate (*P*=0.0116). The mean RT and omission error rate largely remained unchanged.

**Conclusion:** Artificial injection of tACS at 40 Hz over right FP network may improve attention function, especially in the domains of consistency in performance, target/non-target discrimination, and inhibitory control.

## Introduction

Cortical oscillations at various frequencies have been linked to diverse cognitive functions. For instance, working memory and semantic memory functions have been respectively associated with power changes in the theta and upper alpha bands (Klimesch, 1999, Brzezicka et al., 2019). Gamma rhythms have been associated with the functioning of extensive brain networks and cognitive processes such as attention, perceptual grouping, memory, and working memory (Fell et al., 2002, Herrmann et al., 2010, Brookes et al., 2011, Lundqvist et al., 2018). With the introduction of transcranial alternating current stimulation (tACS), it becomes possible to inject artificial oscillation into the brain and study its causal influence on neural rhythmicity and cognition.

Correspondent with neuroanatomical understanding, tACS at theta over anterior frontal and medial temporal and tACS at gamma over occipitoparietal regions lead to improvement in working memory task performance (Park et al., 2022, Manippa et al., 2024a, Wischnewski et al., 2024). The tACS at theta/gamma ranges with anterior/posterior montages may improve episodic memory (Varastegan et al., 2023), while the theta-gamma cross-frequency tACS targeting the prefrontal cortex may enhance visuomotor learning (Diedrich et al., 2024). The tACS at gamma frequency over right parietal and left temporo-parietal regions may respectively modulate endogenous attention and auditory spatial attention (Hopfinger et al., 2017, Wostmann et al., 2018). The promising potential of gamma tACS in addressing mild cognitive impairment (MCI) and early stages of Alzheimer’s disease (AD) has been suggested (Manippa et al., 2024b). Positive evidence regarding the application of tACS to other neuropsychiatric conditions, such as dyslexia, anxiety, depression, and attention-deficit/hyperactivity disorder (ADHD), is beginning to accumulate (Dallmer-Zerbe et al., 2020, Lee et al., 2023).

The clinical effects of tACS appear to be frequency- and region-specific as briefly summarized above. It is noteworthy, however, that sometimes tACS may be detrimental to neuropsychological performance. For example, prefrontal gamma modulation may impair working memory (Wischnewski et al., 2024), and beta tACS may reduce motor cortex excitability (Zaghi et al., 2010). Our recent report suggested that tACS may desynchronize underlying neural oscillation (Lee and Tramontano, 2023). It is thus imperative to empirically examine if a specific tACS protocol is beneficial. It is acknowledged that attention function engages broad neural substrates, relatively right-lateralized for visuospatial processing, particularly fronto-parietal (FP) network (Shipp, 2004, Thiebaut de Schotten et al., 2011). To the best of our knowledge, the influence of gamma tACS over right FP regions on the attention function has never been specifically examined. The importance of investigating transcranial electrical stimulation (tES) on attention cannot be overstated, as attention is essential for the effective execution of other cognitive processes. If tACS does indeed influence attention, it would impact the neuropsychological interpretation of pertinent research.

Among the frequency bands of brain waves, we are particularly interested in gamma, given its coupling with downward spectra (delta to beta), co-bursts with beta in volitional control, close relevance to attention, and its roles in cognitive impairment in various neurocognitive conditions (Fell et al., 2002, Fan et al., 2007, Foster and Parvizi, 2012, Goodman et al., 2018). This study was designed to trace the changes in attention function via the Test of Variables of Attention (TOVA) following 12 sessions of tACS treatment (Leark et al., 2008).

## Materials and Methods

### Participants

The research protocol centered on patients exhibiting attention and other cognitive deficits who underwent 40 Hz tACS treatment applied to the right frontal and parietal regions. A consent form was acquired for each patient before the commencement of treatment. We conducted a review of data collected from our clinics between 2018 and 2022, with prior approval from a private Review Board (Pearl IRB; https://www.pearlirb.com/). Forty-four cognitively impaired patients were identified who underwent full tACS treatment as well as pre- and post-treatment TOVA assessments.

### Administration of tACS

We employed the FDA-approved neuromodulation device called Starstim-8, developed by Neuroelectrics, Inc., to address cognitive deficits arising from different causes. The device utilizes electrodes connected through wires to a rechargeable battery, delivering electric currents directly to the scalp and brain. Prior to attaching the electrodes, the scalp underwent a gentle cleansing with skin preparation gel. Subsequently, conductive gel was applied to facilitate proper electrode-to-scalp contact. These steps were essential to maintain optimal electrode contact and ensure that the impedance level remained below 5 k ohms (DaSilva et al., 2011).

The montage of electrodes covered the right lateral side of the head at F4 and P4 positions in terms of the 10-20 EEG convention. The peak current intensity was 2.0 mA, with sinewave currents oscillating at 40 Hz and alternating between electrodes F4 and P4, see Figure 1 (A). The simulation of electric field distribution is illustrated in Figure 1 (B). During the neuromodulation session, the subjects were asked to practice computerized cognitive training games (https://www.happyneuronpro.com/). To familiarize patients with tES, we implemented a gradual dose escalation strategy during the initial three sessions. This strategy involved the following current levels: 1.0 mA for the first session, 1.5 mA for the second session, and 2.0 mA for the third session. Each stimulation session had a duration of 20 minutes, commencing with a 1-minute ramp-up phase and concluding with a 30-second ramp-down phase to minimize skin irritation. The 12 treatment sessions were completed in 3 weeks.

**Fig. 1.**
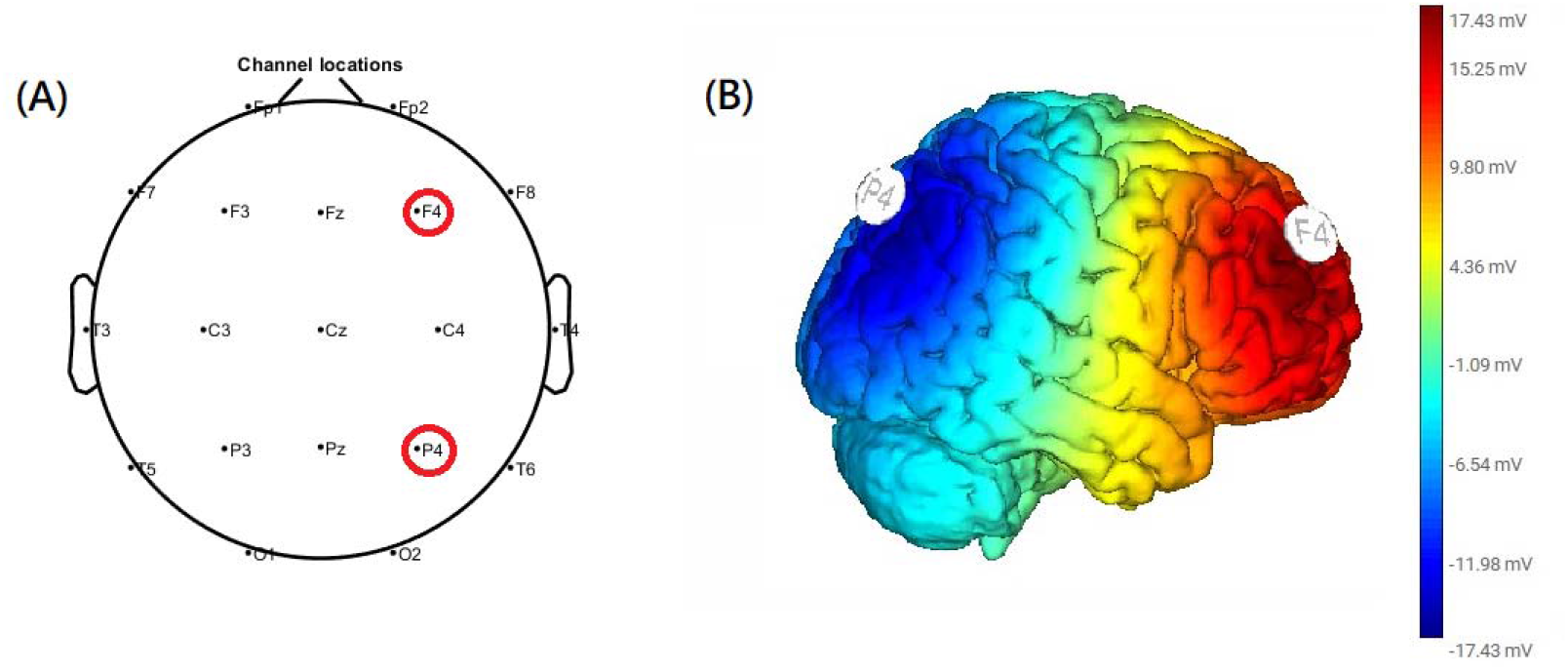
(A) An illustration of the F4 and P4 positions on the scalp. (B) Simulation of tACS at 40 Hz over right FP regions.

### TOVA administration

Before the initial treatment and after the last neuromodulation session, TOVA was administered via the platform Inquisit (https://www.millisecond.com/). TOVA is an individually administered computerized test designed to evaluate attention and impulse control in both normal and clinical populations (Leark et al., 2008). This study exclusively reported findings related to the visual mode, although acknowledging the availability of both visual and auditory modes for assessment purposes. The visual TOVA presents two easily distinguishable geometric figures (target and non-target) at the center of the computer screen. These stimuli appear for 100 milliseconds at intervals of 2000 milliseconds. Participants are instructed to respond to the target stimulus as rapidly as possible. During the first half of the test (stimulus infrequent condition), the target stimulus is presented in 22.5% of the trials (n=72), whereas during the second half (stimulus frequent condition), it appears in 77.5% of the trials (n=252). By manipulating the ratio of target to non-target stimuli, the design allows investigating the impact of varying response demands on the performance. Lumping together all the trials, this preliminary report focuses on five overarching indices: mean reaction time (RT), variability of RT, d-Prime (an indicator of the discriminability between target and non-target), commission errors, and omission errors. The TOVA manual provides age- and gender-matched norms (mean and standard deviation) so that the severity of impairment before treatment can be appraised by the converted z-scores.

## Results

The age of the 44 patients ranged from 6.2 to 70.3 years, with a mean ± SD of 25.1 ± 17.6 years. Among them, there were 28 males and 16 females. All patients tolerated the maximum tES current at 2.0 mA. Their diagnoses include ADHD (n=13), mild to moderate intellectual disability (9), learning disability (4), autism (3), non-amnestic MCI (9), AD-MCI (2), traumatic brain injury (2), and mood disorder (2). At baseline, the average RT variability and d-Prime are worse than age-matched norms by z-scores 3.3 and 2.3. Commission and omission errors are also higher than normal population (z-scores are absent due to deviations of the raw scores from a normal distribution). The side effects were minimal, consistent with another tACS study we conducted (Lee et al., 2023).

**Table 1.**
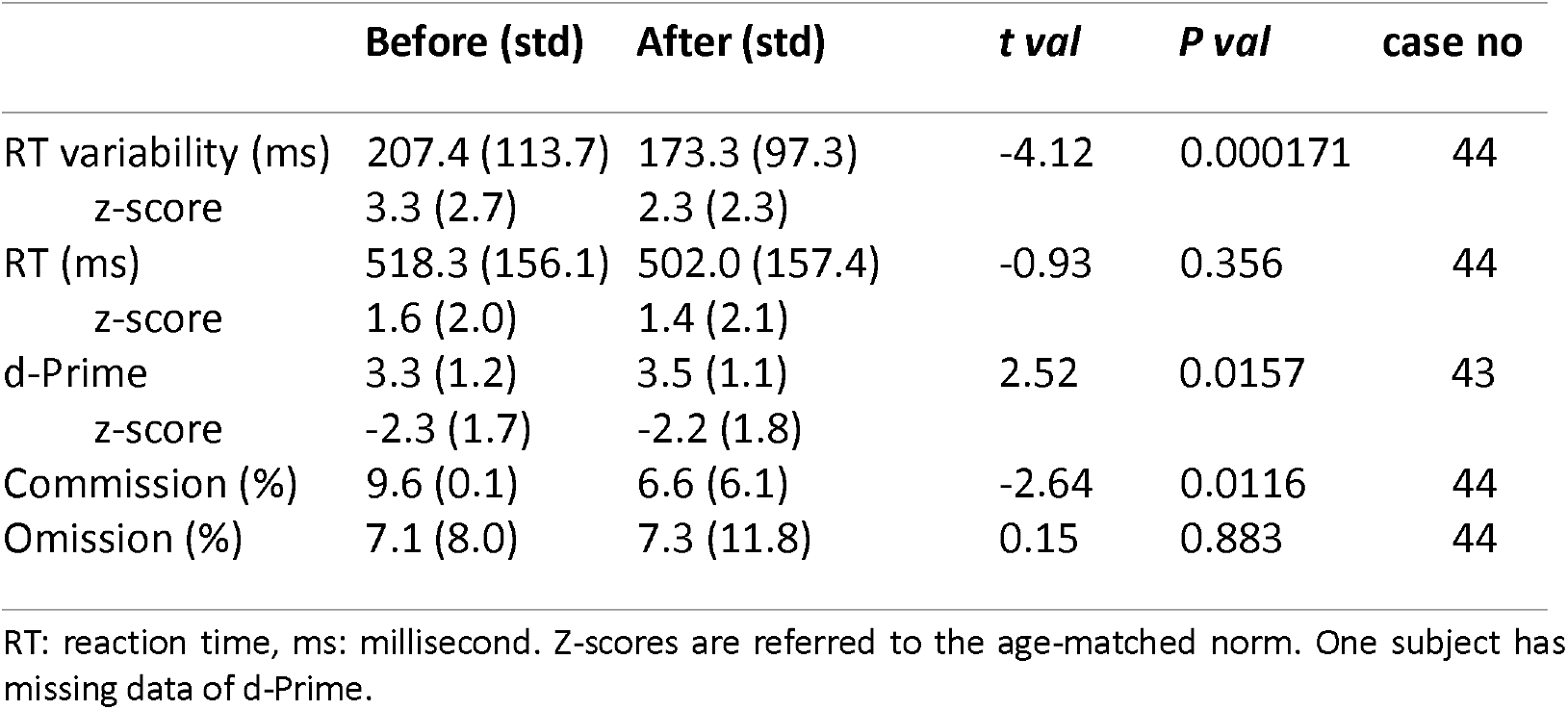
Paired t-tests of TOVA profiles before and after gamma tACS treatment.

|  | Before (std) | After (std) | t val | P val | case no |
| --- | --- | --- | --- | --- | --- |
| RT variability (ms) | 207.4 (113.7) | 173.3 (97.3) | -4.12 | 0.000171 | 44 |
| z-score | 3.3 (2.7) | 2.3 (2.3) |  |  |  |
| RT (ms) | 518.3 (156.1) | 502.0 (157.4) | -0.93 | 0.356 | 44 |
| z-score | 1.6 (2.0) | 1.4 (2.1) |  |  |  |
| d-Prime | 3.3 (1.2) | 3.5 (1.1) | 2.52 | 0.0157 | 43 |
| z-score | -2.3 (1.7) | -2.2 (1.8) |  |  |  |
| Commission (%) | 9.6 (0.1) | 6.6 (6.1) | -2.64 | 0.0116 | 44 |
| Omission (%) | 7.1 (8.0) | 7.3 (11.8) | 0.15 | 0.883 | 44 |

RT: reaction time, ms: millisecond. Z-scores are referred to the age-matched norm. One subject has missing data of d-Prime.

## Discussion

tES has been widely applied as a tool for basic and clinical research (Guleyupoglu et al., 2013). In addition to offering a research gateway to modulate brain rhythms and cognitive processes, studies have begun to support tACS as a treatment option for neuropsychiatric conditions (Lee et al., 2023, Manippa et al., 2024b). Previous research has investigated the potential of tACS at the gamma range to ameliorate various cognitive dysfunctions, predominantly episodic memory and working memory (Park et al., 2022, Varastegan et al., 2023, Manippa et al., 2024a, Wischnewski et al., 2024). Relatively few studies have delved into its impact on attention function (Hopfinger et al., 2017, Wostmann et al., 2018). Using TOVA, we evaluated attention function changes subsequent to 12 treatment sessions of 2mA tACS at 40 Hz over right frontal and parietal regions, aiming to ameliorate cognitive deficits stemming from diverse diagnoses. Through the comparisons of post-treatment minus baseline TOVA indices, our results showed that the treatment protocol benefited several, not all, aspects of attention function, including reduction of variability in RT, decrease of commission error, and improvement in separability of target and non-target, i.e., d-Prime. The mean RT and omission error rate largely remained unchanged.

Each of the five selected indices from TOVA addresses a different domain of attention function (Leark et al., 2007). RT provides a general measure of processing speed and efficiency, whereas the variability of RT indicates sustained attention or the stability of performance. Commission and omission error rates respectively reflect impulsivity (tendency to respond to non-target stimuli) and attentional lapses (failure to respond to target stimuli). The former is relevant to deficits in response inhibition or inhibitory control, and the latter to deficits in vigilance or sustained attention. Overall, our results implied that gamma tACS over right FP network may benefit the consistency in performance and inhibitory control but has little impact on processing speed and attentional lapses. It is not surprising that the mean RT was resistant to the impact of tACS since the task instruction requires the participants to respond as quickly as possible (p.3 in professional manual (Leark et al., 2008)). The improvement in discriminating between targets and non-targets may be more closely linked to the decrease in commission errors rather than omission errors, which appeared to be unaffected by the treatment.

Complicated neural substrates participate in the attention functioning. For example, previous functional MRI research revealed that greater pre-stimulus brain activities in the default-mode network were associated with longer RT, and the BOLD responses in the posterior cingulate, left inferior frontal gyrus, and left middle temporal gyrus increased proportionally with RT (Tam et al., 2015). In contrast, RT variability (intra-individual, as in TOVA) was predicted by the activity in the left anterior cingulate cortex (Johnson et al., 2015). Error-related processing involved a broad neural matrix, including anterior cingulate cortex, pre-supplementary motor area, bilateral insula, thalamus, and inferior parietal lobule (Hester et al., 2004). The neural mechanisms behind the therapeutic benefits of tACS are still being investigated. Nevertheless, with tES initiating low-resolution and wide-ranging neuromodulation (see Figure 1), we posit that our tACS targeting the FP network may influence extensive neural substrates beyond those underneath F4 and P4, thereby modulating neural networks/nodes related to attentional function.

Our research samples were drawn from a heterogeneous mix of neuropsychiatric populations. In conjunction with sporadic reports supporting the efficacy of gamma tACS in enhancing attention (Hopfinger et al., 2017, Wostmann et al., 2018), we propose that gamma tACS may serve as a potential therapeutic intervention for a wide range of neuropsychiatric conditions, irrespective of specific diagnoses. Furthermore, attention, a fundamental cognitive capacity, enables individuals to allocate resources and maintain focus on cognitive tasks to facilitate various cognitive functions. This study offers a new caveat for research on the use of gamma tACS to improve cognitive impairment, prompting consideration of whether the cognitive enhancement observed with tACS treatment is attributable to improved attention. We acknowledged that the results of this preliminary report could be contaminated by practice effects. Double-blind and sham-controlled research is warranted to verify its efficacy and to clarify its role as an adjunct treatment for various conditions of attention and cognitive impairment.

## Data Availability

All data produced in the present study are available upon reasonable request to the authors

## Authors Contributions

All authors contributed intellectually to this work. TW Lee and A Sergio carried out the analysis and wrote the first draft together. All authors revised and approved the final version of the manuscript.

## Acknowledgments

This work was supported by NeuroCognitive Institute (NCI) and NCI Clinical Research Foundation Inc.

## Ethical statement

This IRB-approved research analyzed the databank collected from 2018 to 2022. The authors assert that all procedures contributing to this work comply with the ethical standards of the relevant national and institutional committees on human experimentation and with the Helsinki Declaration of 1975, as revised in 2008.

